# Correlation of ELISA based with random access serologic immunoassays for identifying adaptive immune response to SARS-CoV-2

**DOI:** 10.1101/2020.07.06.20145938

**Authors:** Nguyen N. Nguyen, Manohar B. Mutnal, Richard R. Gomez, Huy N. Pham, Lam T. Nguyen, William Koss, Arundhati Rao, Alejandro C. Arroliga, Liping Wang, Dapeng Wang, Yinan Hua, Priscilla R. Powell, Li Chen, Colin C. McCormack, Walter J. Linz, Amin A. Mohammad

## Abstract

Public health emergency of SARS-CoV-2 has facilitated diagnostic testing as a related medical countermeasure against COVID-19 outbreak. Numerous serologic antibody tests have become available through an expedited federal emergency use only process. This paper highlights the analytical characteristic of an ELISA based assay by AnshLabs and three random access immunoassay (RAIA) by DiaSorin, Roche, and Abbott that have been approved for emergency use authorization (EUA), at a tertiary academic center in a low disease-prevalence area. The AnshLabs gave higher estimates of sero-prevalence, over the three RAIA methods. For positive results, AnshLabs had 93.3% and 100% concordance with DiaSorin or Abbott and Roche respectively. For negative results, AnshLabs had 69.7% and 73.0% concordance with DiaSorin and Roche or Abbott respectively. All discrepant samples that were positive by AnshLabs and negative by RAIA tested positive by all-in-one step SARS-CoV-2 Total (COV2T) assay performed on the automated Siemens Advia Centaur XPT analyzer. None of these methods, however, are useful in early diagnosis of SARS-CoV-2.

## Introduction

The SARS-CoV-2 virus outbreak that began in late 2019 in Wuhan, has a mortality rate of approximately 6.1% worldwide [1-3]. Diagnostic testing is necessary for identifying and isolating infected individuals to limit spread of disease. Molecular testing such as reverse-transcriptase polymerase chain reaction (rtPCR) detects active infection; and serology testing helps identify those who were previously infected (including asymptomatic infections) and have recovered [4, 5]. Nucleic acid detection using rtPCR has become the confirmation test, due to its 99% specificity and 60-90% sensitivity within 7 days of exposure [6] but is faced with numerous supply challenges [7]. The United States Food and Drug Administration (FDA) issued an Emergency Use Authorization (EUA) approval for antibody testing as complementary to rtPCR, leading to an explosion of new antibody methods, including rapid diagnostic test (RDT), enzyme-linked immunosorbent assay (ELISA), virus neutralization assay (VNA), and chemiluminescent immunoassay (CLIA). These methods offer a range of sensitivities; the RDT provides results in less than 30 min for the presence or absence of antibodies against the virus in a whole blood specimen but has the lowest sensitivity, ELISA and CLIA can quantify antibodies to the virus in about 2-5 hours and 0.5-1 hour respectively in either serum or plasma; while VNA can quantify presence of active antibodies that are able to inhibit virus growth ex vivo, but requires 3-5 days [8, 9]. The best clinical utility of antibody testing for efficient diagnosis at tertiary medical centers remains unclear for screening asymptomatic patients and is being considered for identifying patients with adaptive immune responses for convalescent plasma donor program, or for treating re-positive cases [10]. Additionally the relative performance of many of these assays remains unclear.

We evaluated the performance of COVID-19 serology testing on three random access immunoassay analyzers (RAIA) that are typically found in clinical laboratory across US -Architect i2000 (Abbott Laboratories, Chicago IL), Cobas e601 (Roche Laboratories, Indianapolis, IN), and Liaison XL (DiaSorin, Stillwater, MN) – comparing their performance to an ELISA assay (AnshLabs, Webster, TX) and rtPCR test (Luminex Corporation, Austin, TX). The ELISA microtiter plate-based immunoassay, was automated on Dynex DSX instrument (*Dynex Technologies*, Chantilly, VA, USA) for testing IgG and IgM in serum or plasma.

## Materials and methods

### Specimen Selection

This project used 167 left-over and de-identified human serum specimens collected and stored at –20^0^C. This included patients who were either hospitalized with a confirmed COVID-19 diagnosis, seen in the Emergency Department with symptoms for COVID-19, or were screened for COVID-19 before an elective surgery procedure. Fifteen of the 167 samples were from patients that tested positive by rtPCR with a confirmed COVID-19 clinical diagnosis. These samples were drawn >13 days after rtPCR testing. One hundred and fifty-two serum samples were from patients who tested negative by rtPCR, 134 of these were collected on same day as rtPCR testing. For the remaining 18 samples, the interval between rtPCR and sample collection ranged from 1–48 days. To avoid degradation, the specimens were tested by four methodologies within 12-20 h of each other. Only samples having sufficient serum volume and rtPCR test results were included in the evaluation project.

### Instrumentation and analysis

Table 1 summarizes the characteristics of the four serologic assays we investigated.

**Table 1.**
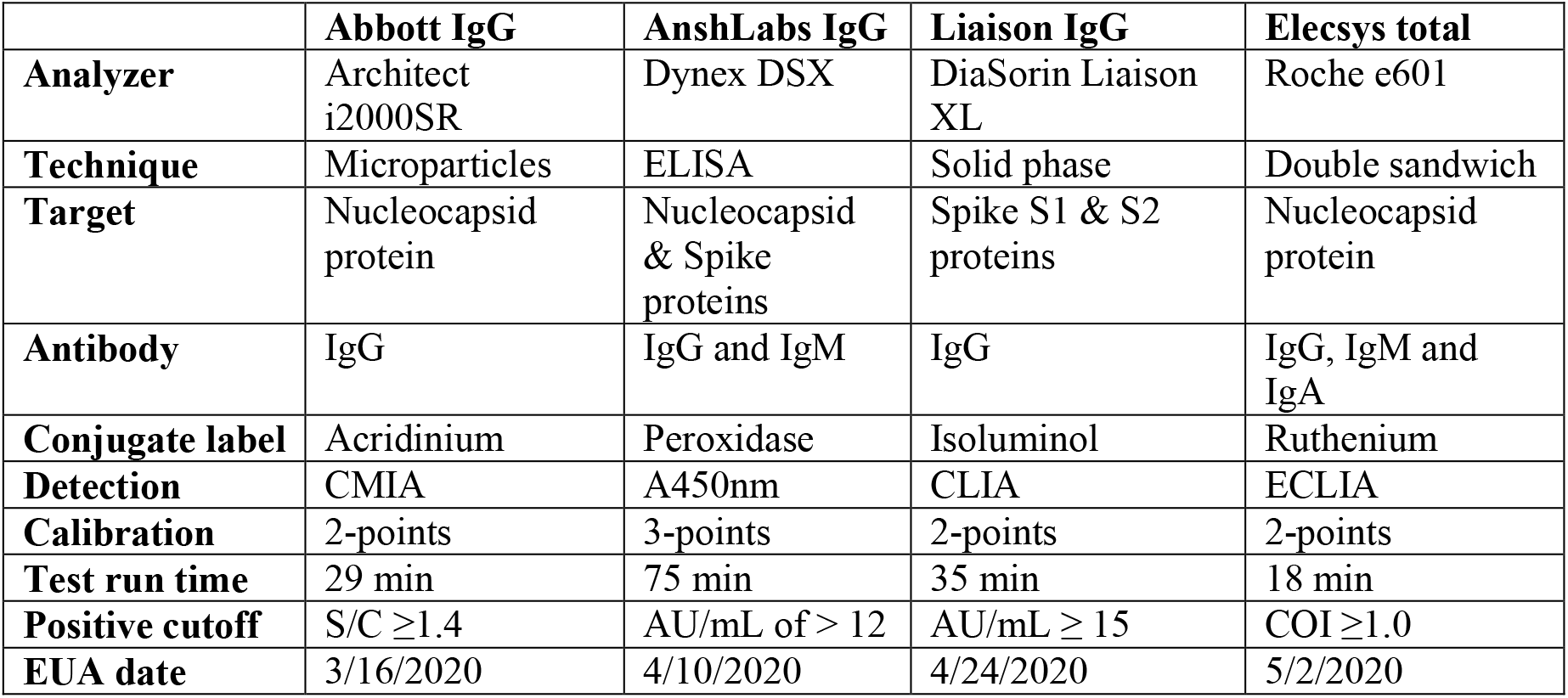
Characteristics summary of four serologic assays. CMIA = chemiluminescent microparticle immunoassay; A450nm = absorbance at wavelength 450 nm; CLIA = chemiluminescent immunoassay; ECIA = Electrochemiluminescent immunoassay. S/C = sample control index ratio; AU/mL = arbitrary concentration units; COI = cutoff index.

|  | **Abbott IgG** | **AnshLabs IgG** | **Liaison IgG** | **Elecsys total** |
| --- | --- | --- | --- | --- |
| **Analyzer** | Architect i2000SR | Dynex DSX | DiaSorin Liaison XL | Roche e601 |
| **Technique** | Microparticles | ELISA | Solid phase | Double sandwich |
| **Target** | Nucleocapsid protein | Nucleocapsid & Spike proteins | Spike S1 & S2 proteins | Nucleocapsid protein |
| **Antibody** | IgG | IgG and IgM | IgG | IgG, IgM and IgA |
| **Conjugate label** | Acridinium | Peroxidase | Isoluminol | Ruthenium |
| **Detection** | CMIA | A450nm | CLIA | ECLIA |
| **Calibration** | 2-points | 3-points | 2-points | 2-points |
| **Test run time** | 29 min | 75 min | 35 min | 18 min |
| **Positive cutoff** | S/C $\geq 1.4$ | AU/mL of $> 12$ | AU/mL $\geq 15$ | COI $\geq 1.0$ |
| **EUA date** | 3/16/2020 | 4/10/2020 | 4/24/2020 | 5/2/2020 |

The AnshLabs SARS-CoV2 IgG assay is based on the ELISA technique that measures antibodies to spike and nucleocapsid proteins. It is for in-vitro diagnostic use only and is performed on the Dynex automated analyzer. Serum samples are diluted in a culture tube and transferred to the microtitration wells coated with purified SARS-CoV-2 recombinant antigen. They are incubated for 30 min at 37°C along with calibrators. The wells are washed and treated with the anti-human IgG antibodies conjugate labeled with peroxidase. After a second incubation and washing step, the wells are incubated with the substrate tetramethylbenzidine (TMB) chromogen solution to induce color change. An acidic stopping solution is added and the degree of enzymatic turnover of the substrate is determined by wavelength absorbance measurement, with 450 nm as the primary filter and 630 nm as the reference filter. The intensity of color change corresponds to arbitrary units of antibody-antigen complex concentration present in the specimen. The analyzer calculates antibody concentration in arbitrary concentration units (AU/mL). Samples with AU/mL of >12, 10–12, and <10 are considered positive, indeterminate and negative for IgG respectively. It is the only test that uses a three-point calibration curve. The sensitivity and specificity are 95.0% and 98.3% respectively [11].

The Abbott SARS-CoV-2 IgG assay was run on the Abbott Architect i2000SR analyzer that measures IgG antibodies to the nucleocapsid protein. The automated, two-step immunoassay uses chemiluminescent microparticle immunoassay (CMIA) technology for qualitative detection of IgG antibodies in human serum. The sample, SARS-CoV-2 antigen-coated paramagnetic microparticles, and diluent are combined and incubated. The antibodies bind to the antigen-coated microparticles. The mixture is washed and anti-human IgG acridinium-labeled conjugate is added. Following incubation, the pre-trigger is added. The resulting chemiluminescent reaction is measured as a relative light unit (RLU). The presence or absence of IgG antibodies is determined by dividing the sample RLU by the stored calibrator RLU to find the IgG assay index (S/C), with a positive cutoff of ≥1.4. The sensitivity and specificity are 100% and 99.63% respectively at ≥ 14 days post onset of symptoms [12].

The LIAISON SARS-Cov-2 S1/S2 IgG is a chemiluminescent immunoassay (CLIA) for detection of anti-S1 and anti-S2 spike glycoprotein specific to SARS-CoV-2 in human serum or plasma on the DiaSorin XL analyzer (Stillwater, MN). Specimen, calibrator, control, coated magnetic particles and diluent are incubated in reaction cuvettes. The antibodies bind to the solid phase through the recombinant S1 and S2 antigens. A second incubation links recombinant S1 and S2 antigens to an isoluminol-antibody conjugate. The starter reagents are then added, and a flash chemiluminescence reaction induced. The light signal, and hence the amount of isoluminol-antibody conjugate, is measured by a photomultiplier and result converted to arbitrary concentration, AU/mL. Samples with AU/mL of ≥15 are considered positive for IgG antibodies. The sensitivity and specificity are 90-97% and 98% respectively ≥ 14 days post onset of symptoms [13].

The Elecsys Anti-SARS-CoV-2 assay is performed on the Roche cobas e601 analyzer for total antibodies specific for IgG, IgM and IgA which target nucleocapsid protein, in human serum or plasma. A 20uL sample and biotinylated SARS-CoV-2 specific recombinant antigen labeled with ruthenium bind in the first incubation. In the second incubation, streptavidin-coated solid phase microparticles are added to help bind the complex to the solid phase via interaction between biotin and streptavidin. The reaction mixture is aspirated into cells where microparticles are captured on the surface of electrode, and the unbound substances are washed out with ProCell solution. The ruthenylated-labeled antigen mediates detection via electrochemiluminescence, which is measured by a photomultiplier tube. Results are calculated by software, comparing the electrochemiluminescence signal of the sample to the cutoff value of the calibration as a cutoff index (COI). Samples with COI ≥1.0 are considered reactive or positive for anti-SARS-COV-2 antibodies. The sensitivity and specificity are 65.5-100% and 99.81% respectively [14].

### Precision and specificity analysis

The precision studies were carried out by testing pooled positive and negative patient specimens for 5 days in duplicate. No discrepant results were noted, i.e. all positive and negative were consistent. The test specificity towards the common cold coronavirus was evaluated by testing 100 prepandemic plasma samples that were collected in October 2019 and stored at −80°C. All samples were from asymptomatic patients who were being evaluated for thyroid disorder.

### Dilution studies

In order to rule out non-specific binding, samples that tested positive by ELISA assay were diluted using sample diluent provided in the AnshLabs assay kit. We made and reran samples for a 1:2, 1:3 and 1:4 dilution, and calculated percent recovery.

### Third party adjudication studies

All ELISA and RAIA discordant result samples were evaluated against the FDA emergency used approved all-in-one step SARS-CoV-2 Total (COV2T) assay performed on the automated Siemens Advia Centaur XPT analyzer.

### Statistical analysis

All test results were collated using a Microsoft Excel (Microsoft, Redmond, WA) spreadsheet. Concordance was calculated using the macro formula in Excel.

## Results

The specificities of the validated in-house AnshLabs SARS-CoV-2-IgG and IgM are listed in Table 2. The cross reactivity to anti-influenza B IgG (5 samples), anti-respiratory syncytial virus IgG (5 samples), anti-nuclear antibodies (5 samples), rheumatoid factors (5 samples), anti-influenza A IgG (5 samples), anti-HCV IgG (5 samples), anti-HBV IgG (5 samples), anti-Haemophilus influenza IgG (5 samples) and anti-HIV (5 samples) was determined by testing 45 patient samples obtained before the pandemic and were positive for these analytes. No cross-reactivity was noted for either SARS-CoV-2-IgG or IgM. The clinical sensitivity and specificity using rtPCR results as the gold standard were found to be 86.7and 91.2% respectively. All samples used for the sensitivity and specificity evaluation were collected from symptomatic patients, either hospitalized inpatients or treated in Emergency Department. The interval between rtPCR confirmation and serology testing ranged from 2-12 days. The specificity toward common cold coronavirus is shown in Table 3. Three of 100 prepandemic samples tested positive for IgG by ELISA and none tested positive by RAIA methods, thereby giving a calculated specificity of 97% and 100% for ELISA and RAIA respectively.

**Table 2.** Specificity of AnshLabs SARS-CoV-2 IgG and IgM assays before and during COVID-19 outbreak for asymptomatic and negative individuals

| Subjects | No of samples | IgG (-) | IgM (-) |
| --- | --- | --- | --- |
| Asymptomatic adults (during COVID-19 outbreak) | 40 | 39/40= 97.5% | 40/40= 100% |
| Presumed negative adults (prepandemic) | 100 | 100/100= 100% | 100/100= 100% |
| Presumed negative pediatric (prepandemic) | 39 | 39/39= 100% | 39/39= 100% |
| **Total** | 179 | 178/179= 99.4% | 179/179= 100% |

**Table 3.**
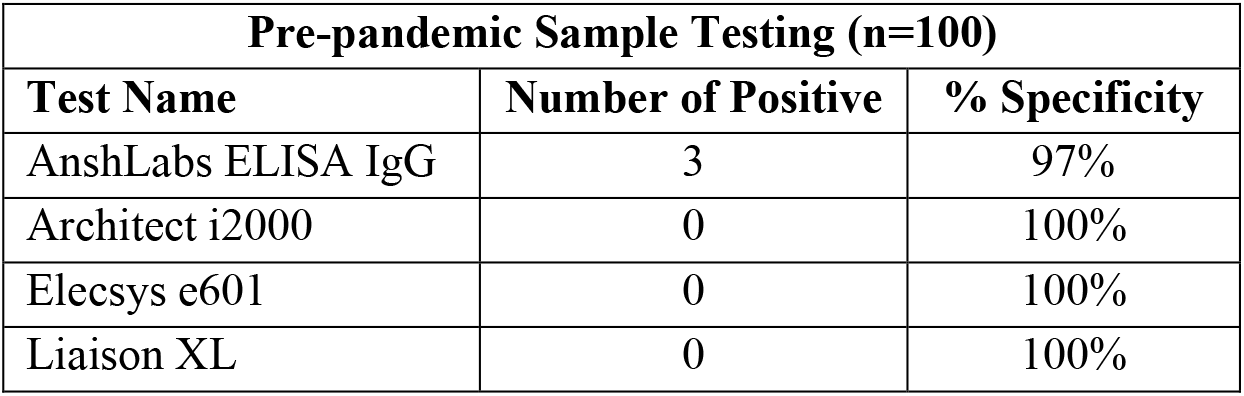
Observed specificities towards common cold coronavirus

Table 4 shows the concordance between ELISA and RAIA results for samples that were confirmed positive for SARS-CoV-2 by rtPCR. These samples were collected from symptomatic patients > 13 days post rtPCR confirmation. ELISA assay correlated best with Total Antibody assay on Roche Elecsys e601 analyzer. This could possibly be attributed to the measurement of IgG antibodies directed towards multiple antigenic proteins (nucleocapsid & spike) by ELISA or measurement of total antibodies (IgG, IgM, and IgA) on Roche Elecsys e601 analyzer.

**Table 4.** Concordance of 15 rtPCR positive samples between a) ELISA and RAIA systems and b) among three RAIA platforms

| Concordance Between ELISA and RAIA for samples from rtPCR positive patients (N = 15) |  |
| --- | --- |
| AnshLabs IgG vs Architect i2000 | 93.3% |
| AnshLabs IgG vs Elecsys e601 | 100% |
| AnshLabs IgG vs Liaison XL | 93.3% |
| **Concordance Among RAIA platforms for samples from rtPCR positive patients (N = 15)** |  |
| Architect i2000 vs Liaison XL | 100% |
| Architect i2000 vs Elecsys e601 | 93.3% |
| Liaison XL vs Elecsys e601 | 93.3% |

Table 5 shows the concordance between ELISA and RAIA for samples from patients that tested negative for SARS-CoV-2 by rtPCR. The ELISA assay showed a concordance ranging from 69.7–73% with different RAIA methodologies: 34, 1, 7, and 5 patients that had tested negative by rtPCR tested positive for antibodies by ELISA, Architect i2000, Liaison XL and Elecsys e601 methodology respectively. All samples that tested positive by ELISA also test positive by Siemens all-in-one step SARS-CoV-2 Total (COV2T) assay Siemens Advia Centaur XPT analyzer. Thus a higher rate of sero-prevalence is observed by ELISA versus RAIA.

**Table 5.** Concordance of 152 rtPCR negative samples between a) ELISA and RAIA systems and b) among four RAIA platforms

|  |  |
| --- | --- |
| **Concordance Between ELISA and RAIA for samples from rtPCR negative patients (n=152)** |  |
| AnshLabs IgG vs Architect i2000 | 73.0% |
| AnshLabs IgG vs Elecsys e601 | 73.0% |
| AnshLabs IgG vs Liaison XL | 69.7% |
| **Concordance Among RAIA platforms for samples from rtPCR negative patients (n=152)** |  |
| Architect i2000 vs Liaison XL | 96.1% |
| Architect i2000 vs Elecsys e601 | 97.4% |
| Liaison XL vs Elecsys e601 | 94.7% |

The concordance of ELISA and RAIA results with rtPCR is shown in Table 6. All patient tested positive by rtPCR also tested positive by ELISA and Elecsys e601 total antibody. Architect i2000 SARS-CoV-2-IgG and Liaison XL were unable to detect antibodies in one sample. All RAIA methodologies showed high correlation with nucleic acid test for patient samples that tested negative by rtPCR, with concordances ranging from 95.39–99.34 %. The ELISA assay on the other hand showed a concordance of only 72.36% for these rtPCR negative samples.

**Table 6.** Concordance of a) serology systems for rtPCR positives confirmed more than 13 days and b) serology systems for all rtPCR negatives

| **a. CONCORDANCE FOR ALL rtPCR POSITIVE SAMPLES**<br>**DRAWN > 13 days after rtPCR result (N=15)** |  |
| --- | --- |
| rtPCR vs ELISA SARS-CoV-2-IgG | 100% |
| rtPCR vs Architect i2000 SARS-CoV-2IgG | 93.3% |
| rtPCR vs Liaison XL SARS-CoV-2 IgG | 93.3% |
| rtPCR vs Elecsys e601 total antibody | 100% |
| **b. CONCORDANCE FOR ALL rtPCR NEGATIVE SAMPLES**<br>**(N=152)** |  |
| rtPCR vs ELISA SARS-CoV-2 IgG | 72.4% |
| rtPCR vs Architect i2000 SARS-CoV-2 IgG | 99.3% |
| rtPCR vs Liaison XL SARS-CoV-2 IgG | 95.4% |
| rtPCR vs Elecsys e601 total antibody | 96.7% |

The non-specific binding dilution data of the AnshLabs assay showed five samples with various concentration levels of IgG were serial diluted to 1:2, 1:4: 1:8 and 1:16. All samples gave a consistent dilution pattern and expected 90-100% recovery of neat sample in AU/mL units (Fig 1).

**Fig 1.**
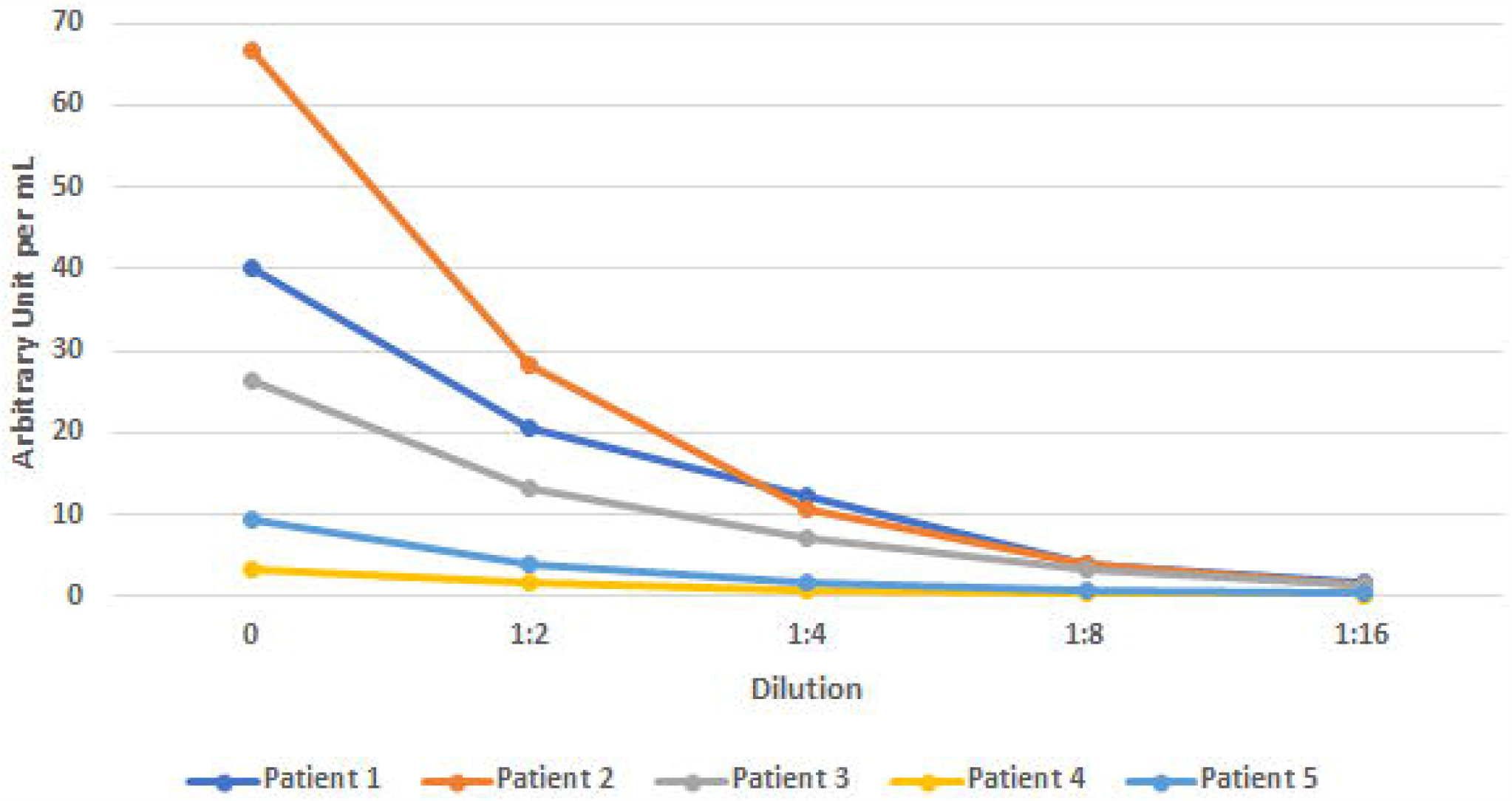
Graph of 5 patient samples diluent sets (1:2, 1:4, 1:8, and 1:16) versus AU/mL levels, ruling out non-specific binding in AnshLabs ELISA assay

## Discussion

All RAIA methods correlated well with ELISA and rtPCR for samples collected >13 days post rtPCR confirmation. There were no significant differences among the methods which tested for IgG targeted to one or both nucleocapsid and spike proteins, or tested for total antibodies.

ELISA detected higher sero-prevalence in rtPCR negative samples than the RAIA methods. This may be due to i) higher analytical sensitivity or a lower cutoff by ELISA, which triggered more positive results; ii) cross reactivity to other coronavirus; iii) non-specific binding of other antibodies, for example autoimmune antibodies or deposition of detection antibody on the microtiter well which led to increased absorbance causing false positives

ELISA assays are generally known for low detection limits in sub ng/mL to low pg/mL because of their increased incubation time thereby allowing antigen-antibody to reach reaction equilibrium and extra washing steps [15, 16]. The Dynex DSX analyzer used for ELISA assay provided optimization flexibility and automation, which is not available on RAIA due to throughput constraint. Cross-reactivity to other coronavirus was evaluated by testing 100 prepandemic samples and found to be 3% and 0% for ELISA and RAIA respectively. The differences in cross-reactivity may account for one or two false positive results, but not for all 34 and 15 positives picked up by ELISA. Non-specific deposition of other antibodies in patient samples or detection antibody was ruled out by dilution studies for ELISA. Recovery of 90–110% ruled out non-specific binding as a possible cause for false positives (Fig 1). The difference in results for positive and negative samples by RAIA methods may also be due to a higher threshold for positivity. The rtPCR assay is used as the gold standard in maximizing analytical sensitivity and specificity during method development which is the most accurate in the early days of the infection when antibody development is low and results in the reported sensitivity of 10-60% on samples collected <14 days post rtPCR confirmation [17-19].

We believe that higher rate of positivity observed for ELISA i.e. 34 versus 1 by Architect, 7 by Liaison XL and 5 by Elecsys e601, is the net effect of extra washing and longer incubation times used by ELISA or a higher S/C cutoff set in RAIA assays. These are not false positives as claimed in other studies [17, 18] but are true positives not picked up by RAIA. This inadvertently decreases identification of infected patients 5-10 days post infection. The recently released all-in-one step SARS-CoV-2 Total (COV2T) assay performed on the automated Siemens RAIA - Advia Centaur XPT analyzer has resolved some of these issues and it correlates well with our in-house ELISA assay by detecting all 34 samples that were missed by other RAIA as positives.

### Project Limitations

Our quality assurance project has some notable limitations. At this stage of the disease, true clinical sensitivity and specificity for different methodologies is difficult to determine because of our limited understanding of the disease process and kinetics. Secondly, our assumption that ELISA has better limits of detection is based on circumstantial evidence, as certified standards quantifying limits of detection on different platforms are not available. Third, the cutoffs provided by manufacturers were relied on which may not have undergone extensive validation. Establishing laboratory specific cut-off is akin to establishing reference ranges, which is highly dependent on prevalence of disease in local population.

## Conclusion

All of the assays we investigated would work well for epidemiological sero-prevalence studies. Among rtPCR negative patients, ELISA gave higher estimates of sero-prevalence in our dataset and would probably do so in population-based epidemiological surveys using serological testing. RAIA methods could however offer other advantages over ELISA which includes i) faster turnaround time; ii) random access to allow immediate testing; iii) longer calibration stability, obviating the need to perform daily calibration as required by ELISA; iv) the ability to perform other immunoassay testing concurrently; and v) higher test throughput and walk away capabilities. However in conclusion, no serological method tested has sensitivity and specificity greater than or equal to 99% for one to 5 days post exposure, limiting their use in early diagnosis.

## Data Availability

Data can be given upon request

## Acknowledgement

The authors would like to thank Ms. Briget Da Graca for editorial comments and manuscript revision. We also acknowledge Ms. Laura Gonazales and her team from Health Texas Provider Network (Dallas, TX) for correlation of AnshLabs results using Siemens Centaur total antibody assay.

